# The Relationship Between an Agricultural Intervention Integrated Within a Group-Based Microfinance Program and Depression and Psychological Stress in Rural Kenya

**DOI:** 10.64898/2026.07.23.26358808

**Authors:** Charles Opondo, James Kamadi, James Akiruga Amisi, Ricky Camplain, Sonak D. Pastakia, Nana Gletsu-Miller, Christina Ludema, Molly Rosenberg

**Affiliations:** Department of Epidemiology and Biostatistics, School of Public Health, Indiana University, 1025 E. 7TH Street, Bloomington, IN, United States; Academic Model Providing Access to Healthcare (AMPATH), P.O. Box 4606, 30100, Eldoret, Kenya; Department of Family Medicine, Moi University School of Medicine, P.O. Box 4606 30100, Eldoret, Kenya; Purdue Kenya Partnership, College of Pharmacy, Purdue University, 640 Eskenazi Ave. Indianapolis, IN 46202,USA; Department of Applied Health Sciences, School of Public Health, Indiana University, 1025 E.7TH Street, Bloomington, IN, USA

**Author notes:** **Corresponding Author:** Charles Opondo; Department of Epidemiology and Biostatistics. School of Public Health, Indiana University, 1025 E. 7^TH^ Street, Bloomington, Indiana, USA.

**Keywords:** Agricultural Intervention, Depression, Psychological Stress

## Abstract

**Background:** Poverty is a social determinant of poor mental health outcomes. Agricultural interventions may reduce the burden of poor mental health outcomes for depression, and stress by enabling economic pathways out of poverty, particularly when paired with a group-based microfinance programs that provide complementary economic support. We estimated the relationship between an agricultural intervention, depression and psychological stress outcomes.

**Methods:** We conducted a cross-sectional survey using validated depression and stress scales among 312 participants. The exposed group received farm inputs and agribusiness trainings. We fit an adjusted multivariate linear regression models to estimate the association for depression and stress scores, in addition to a quantile regression model to examine variation of effect sizes across depression score quantiles. We tested for effect measure modification by wealth status as measured with a composite score derived from 20-household and agricultural assets.

**Results:** The agricultural intervention was associated with lower depression (β = −1.81; 95% CI: −4.24, 0.62) and stress scores (β = −1.39; 95% CI: −3.35, 0.56), but estimates were imprecisely measured. The association was strong in those with highest depression scores (Q90: β = −7.30; 95% CI: −13.59, −0.99), and weaker in those with lower depression scores (Q10: β = −0.7; 95% CI: −2.93, 1.48). There was no effect measure modification by wealth status (Wald p = 0.24), although larger effect sizes for depression were observed in those with low household wealth (β = −5.23; 95% CI: −10.75, 0.33).

**Conclusion:** An agricultural intervention was associated with modestly lower depression and stress scores, although overall estimates were imprecise. The association was strongest among individuals with severe depression, suggesting agricultural support may be beneficial for those experiencing greater psychosocial burden. Integrating agricultural support into microfinance programs may reduce depression among individuals with greater need. Future longitudinal work with larger samples may improve our causal understanding of these relationships.

## Introduction

Depression and psychological stress are the leading causes of morbidity globally, affecting an estimated 332 million people ^1^. In sub-Saharan Africa, approximately 29 million people experience symptoms related to depression and stress^2^. Approximately 2 million people experience either depression or stress-related symptoms in Kenya ^33,4^ . People with depression and stress are at higher risk of multiple morbidities such as cardiovascular diseases and premature mortality ^5–7^ , while early depressive symptoms strongly predict the development of future depression ^8^ .

While the precise cause of depression and stress remain unclear, current evidence suggests a complex interaction of biological, genetic, environmental, and social risk factors ^9–11^ . A consistent social risk factor for depression and psychological stress is poverty. Poverty may affect mental health through food insecurity. Food insecurity can increase psychological stress associated with uncertainty about consistent access and availability of food as well as lack of essential nutrients critical for preventing depression ^12,13^ . Thus, livelihood interventions that promote economic strengthening may reduce the burden of psychological stress and depression thereby improving the psychological well-being of people living in poverty.

Agricultural interventions are structured livelihood programs designed to promote sustainable farming practices, improve food production, and/or enhance nutritional outcomes through the provision of farm input support, training and linkage to markets for sale of agricultural produce. Agricultural interventions may have the potential to reduce depression and psychological stress outcomes through the following plausible pathways. First, agricultural interventions may increase household income and purchasing power while also strengthening social connections through program participation. Together, these factors may prevent psychological stress associated with difficulty in access to consistent availability of food thereby contributing to improved mental well-being ^14^ . Second, participating in agricultural interventions may reduce depression through a nutritional pathway by improving the availability and consumption of different food groups ^15^ , thereby reducing deficiencies in key micro- and macro-nutrient deficiencies that are associated with increasing the risk of depression ^16,17^ .

Despite the promise of agricultural interventions, the existing evidence base on their association with depression, and psychological stress outcomes is limited in scope, study design and generalizability. In particular, agricultural interventions in Kenya and Tanzania were associated with reductions in anxiety, psychological stress and depression outcomes, but findings from these studies may not be generalizable to broader populations as they were focused only among people living with HIV ^14, 18 19–21^ . In contrast, prior work on more holistic economic interventions in South Africa,^22^ Kenya,^23^ Mexico,^24^ Tanzania,^25^ Brazil,^26^ and microfinance programs in sub-Saharan Africa, and Asia ^27–29, 30,31^ , are mixed. The mixed scientific evidence base reflects contextual differences and variation in intervention design, which limit the generalizability of prior findings to other settings. While limited and mixed, prior work evaluating the associations between agricultural or broader socioeconomic interventions and depression outcome have rarely assessed effect measure modification by wealth status leaving uncertainty as to whether these interventions benefit individuals differently across socioeconomic strata. This highlights an important research gap regarding the association between an agricultural intervention embedded within a group-based microfinance program and depression and psychological stress outcomes in Western Kenya.

In this study, we aimed to estimate the association between exposure to an agricultural intervention embedded within a microfinance program and depression and psychological stress outcomes in rural Western Kenya. We hypothesized that participation in the agricultural intervention would be associated with lower prevalence of depression and psychological stress compared to participants who did not receive the intervention and that the estimated association would be modified by household wealth.

## Materials and methods

### Study Design and Setting

We conducted a cross-sectional study between June and August 2025 among <u>B</u>ridging Income <u>G</u>eneration through gro<u>uP I</u>ntegrated <u>C</u>are (BIGPIC) microfinance program participants in the village wards of Matulo and Milo in Webuye sub-County of Bungoma County in rural Western Kenya. The two wards are similar in sociodemographic makeup and a population size (∼15,000 residents/ward). Bungoma County is characterised by a high poverty rate at 36% which is slightly above the national poverty rate of 32% ^32^ . Agriculture is the main economic activity in Matulo and Milo wards, although some residents engage in small-scale business activities for household income. The BIGPIC microfinance program has been established and operated since the 2013 in Milo and Matulo ^33,34^ . The BIGPIC program was designed to address the structural barriers to healthcare access and supports socioeconomic enhancement activities through the provision of agricultural interventions and business literacy education to improve the economic well-being of its members. The BIGPIC program is affiliated with the Academic Model Providing Access to Healthcare (AMPATH). AMPATH is a consortium of North American Universities in partnership with Moi University, Moi Teaching and Referral Hospital and the government of Kenya, under the leadership of Indiana University with a goal of improving care, training providers, and conducting locally relevant research.

The agricultural intervention within BIGPIC program includes training in sustainable farming practices, poultry and livestock rearing, cultivation of African indigenous vegetables, provision of farm inputs, and linkage to markets for the sale of agricultural produce ^35,36^ .

### Study Population

Based on participants’ exposure status to the agricultural intervention, we purposively recruited 312 eligible participants from a roster of 4545 BIGPIC program members. We identified 156 participants who received the agricultural intervention (exposed) and 156 who did not receive the agricultural intervention (unexposed). Participants were eligible to participate if they were members of the BIGPIC microfinance program, a resident of Matulo or Milo wards for a least one year prior to data collection and aged 18 years and above.

### Data Collection

Interviewer-administered surveys were conducted using REDCap ^37^ . Three trained research assistants administered surveys to eligible participants during weekly group meetings in a privately secured location within the homesteads to ensure confidentiality.

### Ethical Consideration

Two ethical approvals were obtained from (1) Indiana University’s Institutional Review Board (#25993) in the United States of America and (2) Moi University’s Institutional Research and Ethics Committee (#0005080) in Kenya. We additionally obtained permission to conduct the study from the Kenya National Commission for Science, Technology and Innovation (NACOSTI, #P/25/4175009). Informed consent forms were translated into the local dialect (‘Kibukusu’) before administration. Prior to administration of the survey, written and verbal informed consent was obtained from all participants, while verbal informed consent was witnessed and formally recorded as per the guidelines laid down in the Declaration of Helsinki ^38^ .

### Key Variables Primary exposure

The primary exposure variable was the agricultural intervention. Exposure to the agricultural intervention was defined as the receipt of one cycle of farm input support, twenty-eight cycles of trainings on land preparation, nursery seedbed preparation for African indigenous vegetables, livestock, poultry rearing and linkage to markets for the sale of agricultural produce. We measured participation in the agricultural intervention using BIGPIC program administrative records provided by the BIGPIC program leaders. We verified exposure status by asking participants about their previous participation in agricultural training and receipt of subsidized farm input supplies. Participants who reported themselves as having been exposed to the agricultural intervention but had their exposure information missing from BIGPIC program administrative records were excluded from the survey. Participants unexposed to the agricultural intervention were newly enrolled members of the BIGPIC program seven months prior to the data collection period and were recruited to the study after verifying their membership status from the records provided by BIGPIC program group leaders.

### Primary Outcomes

We measured depression using the Centre for Epidemiologic Studies for Depression (CES-D) scale ^3940^ . The CES-D scale was selected based on evidence of its prior validation across four sub-Saharan African countries including Kenya, in a community-based, low-resource settings with socioeconomic and demographic characteristics similar to those of our study sample. In a population comparable to our study sample, the CES-D demonstrated a good internal consistency with Cronbach’s α ranging from 0.85 to 0.90 ^41^, supporting the reliability of the scale in a setting similar to the current study. Participants reported experiencing depressive symptoms over the past seven days. Responses were recorded using a 4-point Likert scale (0 = rarely or never, 1 = sometimes and 2 = often, 3 = most of the time). Items assessed the frequency of symptoms such as “I had trouble keeping my mind on what I was doing,” “I felt hopeful about the future,” and “I felt my life had been a total failure,” among others on the scale. Four positively worded questions were reverse coded, and responses were summed resulting in a total depression score range from 0 to 53. Higher depression scores was suggestive of probable severe depression.

Psychological stress was assessed using the 10-item Perceived Stress Scale (PSS-10), a validated instrument that has been used previously in sub-Saharan African settings ^42–44^ . The PSS-10 has not been formally validated in a Kenyan population. However, the scale has been validated in a rural and low-resource settings with socioeconomic and demographic make-up that is comparable to our study population, demonstrating a good internal consistency of Cronbach’s α = 0.64-0.93 ^42^. We queried participants about their 10 stress-related experiences over the past one month using a 5-point Likert scale (“0 = never”, to “4 = very often”). Four positively worded items were reverse coded, and responses were summed to generate total psychological stress scores. Total PSS-10 scores ranged from 0 to 40 and were categorized for descriptive purposes as follows: scores of 0–13 indicated low stress, 14–26 indicated moderate stress, and 27–40 indicated a probable higher stress levels. Because these categories are intended for descriptive interpretation only ^45^ , and do not represent clinical cutpoints ^46^ , we treated psychological stress as a continuous outcome variable in a multivariate analysis to determine its relationship with the agricultural intervention.

### Covariates

We also collected data on sociodemographic characteristics to contextualize our study and address potential confounding: age, calculated as the number of years from birthdate, educational attainment, measured as number of years of schooling completed, marital status, self-reported as married, never married, widowed, divorced or separated, household size measured as the number of people living in the same household; sex, self-reported as either male or female, work status as employment outside the homestead during the previous month, and wealth status, calculated using a weighted index derived from a composite score of 20 household and agricultural asset ownership and categorized into wealth quantiles as per the 2022 Kenya Demographic and Health Survey guidelines ^47^ .

## Statistical Analysis

We fit multivariate linear regression models to estimate the association between exposure to the agricultural intervention and each of two outcomes for depression and stress scores. We stratified wealth status as high or low based on values above or below the median wealth index, and examined whether wealth status functioned as an effect measure modifier for the association between the agricultural intervention and depression scores, by including an interaction term between the agricultural intervention indicator variable and wealth status. We controlled for potential confounding in multivariate linear regression models, using a minimally sufficient covariate set identified from the directed acyclic graph (supplementary Figure 1), which included marital status, educational attainment, age, and sex. In addition, we implemented Inverse Probability of Treatment Weighting (IPTW) as a complementary approach to estimate the average treatment effect comparing if the entire study population was exposed to the agricultural intervention versus not exposed. The IPT weights were derived from a logistic regression model estimating the probability of receiving the agricultural intervention using the same minimally sufficient covariate set. We then applied these weights in the overall multivariate linear regression models.

We also fit a quantile regression model to estimate the association between the agricultural intervention and depression across the 10^th^, 25^th^, 50^th^ , 75^th^, and 90^th^ quantiles of the CES-D score distribution. This model was chosen to capture variation in how the agricultural intervention was associated with depression in participants with less and more severe depression symptoms. We then compared the magnitude of effect estimates across quantiles and conducted a statistical test using a joint Wald-type test to assess whether these estimates were significantly different from each other. We conducted data analysis was using RStudio software^48^ .

## Results

### Sample Characteristics

In the overall sample of 312 participants, most were women (87%), and just over half (52%) were aged 30 years or older, with a mean age of 40 years. In the 30 days preceding the survey, 78% reported no paid employment outside their households, and 85% were currently married. Regarding educational attainment, 46% had completed primary education, while 21% and 10% had attained secondary and post-secondary education respectively. The median household size was five members (IQR: 4,7). Sociodemographic characteristics differed between participants who received the agricultural intervention and those who did not. Specifically, individuals who participated in the agricultural intervention were older, married, completed primary education, and belonged to the highest wealth quintile. We did not observe statistically significant differences between the two groups with respect to sex, work status, household size, and depression outcomes (Table 1).

**Table 1.** Study population characteristics in the sample and by agricultural exposure status (n=312)

|  |  |  |  |  |
| --- | --- | --- | --- | --- |
| **Agricultural Intervention Exposure Status** |  |  |  |  |
|  |  | **YES** | **NO** | **p-value** |
| **Sociodemographic characteristics** | **Overall sample (100%)** | **N = 156** | **N=156** |  |
| **Sex** |  |  |  | 0.24 |
| Male | 40 (13%) | 24 (15%) | 16 (10%) |  |
| Female | 272 (87%) | 132 (85%) | 140 (90%) |  |
| **Age Category** |  |  |  |  |
| **<30** | 32 (10%) | 6 (4%) | 26 (17%) | **<0.0001** |
| 30-39 | 106 (34%) | 47 (30%) | 59 (31%) |  |
| 40-59 | 125 (40%) | 69 (44%) | 56 (36%) |  |
| 60 + | 49 (12%) | 34 (22%) | 15 (9%) |  |
| **Marital status** |  |  |  | **0.02** |
| Never married | 46 (15%) | 33(21%) | 13(8%) |  |
| Currently married | 264 (85%) | 122 (79%) | 142(92%) |  |
| Missing | 2 | 1 | 1 |  |
| **Education** |  |  |  | **0.005** |
| None/Some Primary | 67 (21%) | 21 (13%) | 46 (29%) |  |
| Primary | 142 (46%) | 80 (51%) | 66 (42%) |  |
| Secondary | 67 (21%) | 35 (22%) | 32 (20%) |  |
| Post-secondary | 32 (10%) | 20 (12%) | 12 (8%) |  |
| **Worked outside home (last 30 days)** |  |  |  | 0.2 |
| Yes | 69 (22%) | 40 (26%) | 29 (19%) |  |
| No | 243 (78%) | 116 (74%) | 127 (81%) |  |
| **Household size [Median (IQR)]** | 5(4-7) | 5 (4-7) | 5 (4-7) | 0.3 |
| **Wealth quintile** |  |  |  | **0.002** |
| Q1 | 58 (19%) | 22 (14%) | 36 (23%) |  |
| Q2 | 58 (19%) | 21(13%) | 37 (24%) |  |
| Q3 | 81 (26%) | 41 (26%) | 40 (26%) |  |
| Q4 | 97 (31%) | 62 (40%) | 35(22%) |  |
| Missing | 18 | 10 | 8 |  |
| **Depression** |  |  |  | 0.9 |
| Yes | 153 (49%) | 76 (49%) | 77 (49%) |  |
| No | 159 (51%) | 80 (51%) | 79 (51%) |  |

### Agricultural Intervention, Depression and Psychological Stress scores

Overall, people who participated in the agricultural intervention had a trend toward better mental health outcomes than those who did not, though these results were not statistically significant (Table 2). The agricultural intervention participants had almost two point lower depression scores compared to those who did not receive the agricultural intervention (aβ = - 1.81; 95% CI: −4.24, 0.62). The agricultural intervention participants had lower mean stress scores compared to those who did not receive the intervention in the adjusted model (aβ = - 1.39; 95% CI: −3.35, 0.56), though not statistically significant (Table 2).

**Table 2:** Multivariate Linear regression estimates for the association between the agricultural intervention, and depression scores, stratified by wealth status, and stress scores.

|  |  |  |  |  |
| --- | --- | --- | --- | --- |
|  | **Unadjusted**<br>***beta* Estimate**<br>**(95% CI)** | **p-value** | **Adjusted<sup>1</sup>**<br>***beta* Estimate**<br>**(95% CI)** | **p-value** |
| **Depression scores** |  |  |  |  |
| **Full Sample**<br>**n=312** | -2.19 (-4.47, 0.08) | 0.06 | -1.81 (-4.24, 0.62) | 0.14 |
| **Stratified Sample** |  |  |  |  |
| High-wealth n=148 | -0.86 ( -4.82, 3.10) | 0.67 | -0.83 (-5.06, 3.40) | 0.80 |
| Low-wealth<br>n=146 | -4.97 (-9.85, -0.09) | 0.05 | -5.23 (-10.75, 0.33) | 0.06 |
| **Stress scores** |  |  |  |  |
|  | -2.06 ( -3.91, -0.18) | 0.03 | -1.39 (-3.35, 0.56) | 0.16 |

<sup>1</sup>Covariates in adjustment set were marital status, education, age, and sex
95% CI: Confidence interval
P- value for interaction term for the agricultural intervention and wealth status on depression outcome = 0.20

For IPT-weighted results, exposure to the agricultural intervention was associated with over one point lower mean depression score compared to no exposure at the population level (β = − 1.67; 95% CI: −3.94, 0.59 ), although not statistically significant (Supplementary Table 1). In the quantile regression analysis, we found a general trend of stronger effect estimates in people with higher vs. lower depression scores (Table 3). Specifically, among participants who participated in the agricultural intervention, greater reductions in depression scores were observed at 75^th^ percentile (Q75: β = −8.0; 95% CI: −13.9, −2.1), and 90^th^ percentile (Q90: β = − 7.0; 95% CI: −12.7, −1.1), compared to those observed at 10^th^(Q10: β = −1.0; 95% CI: −3.1, 1.1), and 25^th^ (Q25: β = −4.0; 95% CI: −7.3, −0.7) percentiles. After covariate adjustment, the direction of the trend remained but the magnitude of the point estimates were slightly attenuated toward the null, but the larger reductions persisted at the upper tail of the distribution: at 10^th^ (Q10: aβ = −0.7; 95% CI: −2.9, 1.5), 25^th^ (Q25: aβ = −3.5; 95% CI: −6.61, −0.39), at 50^th^ (Q50: aβ = - 1.21,95% CI: −7.82, 5.41), 75^th^ (Q75: aβ = −3.22; 95% CI: −10.18, 3.73), and 90^th^ (Q90: aβ = −7.3; 95% CI: −13.59, −0.99). The results from the statistical test of heterogeneity across quantiles (Wald p-value for Anova joint test = 0.003), indicated that the observed variation across quantiles was unlikely to be due to random variability alone, suggesting that the association between the agricultural intervention and depression scores differed meaningfully across the quantiles.

**Table 3:** Quantile regression estimates for the association between the agricultural intervention and depression scores across various quantiles (n=312)

95% CI: Confidence interval P- value for interaction term for the agricultural intervention and wealth status on depression outcome = 0.20
|  |  |  |  |  |  |  |  |  |  |  |
| --- | --- | --- | --- | --- | --- | --- | --- | --- | --- | --- |
|  | Q10 |  | Q25 |  | Q50 |  | Q75 |  | Q90 |  |
| Depression scores | $\beta$ Estimate<br>(95 CI) | P-value | $\beta$ Estimate<br>(95 CI) | P-value | $\beta$ Estimate<br>(95 CI) | P-value | $\beta$ Estimate<br>(95 CI) | P-value | $\beta$ Estimate<br>(95 CI) | P-value |
| **Unadjusted Model<sup>1</sup>** |  |  |  |  |  |  |  |  |  |  |
| Unexposed group |  |  |  |  |  |  |  |  |  |  |
| Exposed group | -1.00 (-3.08, 1.09) | 0.35 | -4.0 (-7.26, -0.74) | 0.02 | -0.5 (-6.77, 5.78) | 0.88 | 8.04 (-13.92, -2.15) | 0.008 | -7.00 (-13.44, -0.56) | 0.03 |
| **Adjusted Model<sup>2</sup>** |  |  |  |  |  |  |  |  |  |  |
| Unexposed group |  |  |  |  |  |  |  |  |  |  |
| Exposed group | -0.72 (-2.82, 1.38) | 0.5 | -3.50 (-6.60, -0.40) | 0.03 | -1.21 (-7.82, 5.41) | 0.72 | -3.22 (-10.03, 3.59) | 0.35 | -7.29 (-13.63, -0.96 ) | 0.02 |

<sup>1</sup>Covariates in adjustment set were marital status, education, age, and sex <sup>2</sup>Inverse probability of treatment weights were constructed using marital status, education, age, and sex Wald p= value from Anova joint test for heterogeneity of point estimates across quantiles = 0.003 B: Beta coefficient, 95%CI: Confidence Interval.

### Wealth-stratified Analysis

In the wealth-stratified analysis, we found no evidence of effect measure modification by wealth status (Wald p-value for interaction term=0.24; Table 3), likely due to small samples in our study. Nevertheless, the agricultural intervention appeared strongly associated with lower mean depression scores in participants from low- versus high-wealth backgrounds. Among individuals from low-wealth backgrounds, the agricultural intervention was associated with a 5 point lower mean depression scores compared to those who did not participate in the intervention (aβ = −5.23; 95% CI: −10.75, 0.33). Among individuals from high-wealth backgrounds, the agricultural intervention was associated with a much smaller difference in mean depression scores (aβ = −0.83; 95% CI: −5.06, 3.40).

## Discussion

In this study, our findings suggest that agricultural interventions may be associated with fewer depressive symptoms among participants in rural Western Kenya. Beneficial associations with depressive symptoms were particularly strong among individuals with severe depressive symptoms and among participants from low-wealth backgrounds compared with those with mild depressive symptoms and those from high-wealth backgrounds. Overall, these findings suggest that agricultural interventions may be particularly effective in reducing the burden of depression and psychological stress among economically vulnerable populations in low-resource settings like in rural Western Kenya.

Our findings align with prior studies reporting mental health benefits associated with agricultural interventions in low-resource settings ^14,19–21,49^ . In contrast, they differ from prior work that reported no association between cash transfer or microfinance programs, and participants’ mental health outcomes ^25,26,30,31^ . Several factors may explain these contrasts. First, program design and objectives varied substantially across studies. While our study focused on the agricultural intervention embedded within a group-based microfinance program that fostered social connectedness through collective participation, previous cash transfer interventions provided monetary, and economic support to participants, conditional on them fulfilling strict intervention requirements. Second, the potential mental health benefits of traditional microfinance programs may be attenuated by psychological stress associated with loan repayment obligations. In contrast, participants in our study received agricultural training, and subsidized farm inputs without repayment requirements, which may have reduced financial strain and depressive symptoms. Thus, agricultural interventions paired with microfinance programs that provide unconditional economic support and without debt obligations may be more effective for improving mental health than stand-alone cash transfer or traditional microfinance models.

We also observed heterogeneity in the agricultural intervention effects across the distribution of depression scores, and found a stronger association trending in protective direction among participants with more severe depressive symptoms than among those with moderate symptoms. This pattern suggests that mental health benefits’ of agricultural interventions may not be evenly distributed across the populations but instead may be concentrated among individuals with the greatest mental health burden, highlighting their potential value for targeting economically vulnerable and high-risk groups for depression. Another possible explanation for stronger association among those with severe depression is simply that the depression scores are regressing towards the mean, as they tend to converge over time, regardless of any intervention.

We observed stronger associations between the agricultural intervention and lower depression scores in people from low-versus higher-wealth backgrounds. These findings are consistent with an interpretation that high-wealth households may have already had greater baseline economic stability and food security ^50^ . For these individuals, economic stressors may not have contributed as much to their depressive symptoms, meaning an agricultural intervention would be less likely to produce meaningful changes in their psychological well-being. In contrast, participants from low-wealth backgrounds may have experienced substantial gains from agricultural interventions which could translate into larger reductions in depressive symptoms. However, we note that our wealth-stratified estimates should be interpreted cautiously due to small sample sizes and wide confidence intervals, underscoring the need for future work to prioritize larger samples for more precise estimates.

For psychological stress outcome, participants who received the agricultural intervention trended toward experiencing lower mean stress scores, although these findings were not statistically significant. These findings are consistent with prior studies reporting reductions in perceived stress symptoms ^51–53^ . However, the magnitude of the effect estimate observed for psychological stress outcome was smaller than anticipated in our study. The reason why the effect size wasn’t as large as it could have been may be because our study focused primarily on the agricultural intervention designed to improve economic, and livelihood outcomes without an explicit stress-reduction component, unlike prior studies that incorporated mindfulness-and meditation-based components to enhance coping skills and stress regulation.

The potential for confounding was a major concern in our study. We sought to minimize this bias by carefully selecting covariates using the directed acyclic graphs and controlling for them using two complimentary statistical approaches: conventional covariate adjustment and inverse probability of treatment weighting. Given that the two approaches rely on different assumptions to control for confounding, we utilized IPT-weights generated from the same covariates included in multivariate linear regression model to obtain a complimentary point estimate, understanding that IPT-weighting could capture a different estimand derived from multivariate linear regression model. The IPT-weighted results were different, and weaker than the results obtained from multivariate linear regression model, suggesting that higher proportion of exposed participants experienced better mental health outcomes associated with the agricultural intervention. However, unmeasured and residual confounding still remain a threat to the validity of our study findings. Future work should employ randomized controlled trials or quasi-experimental designs to strengthen our confidence in the findings .

The cross-sectional design of our study precludes establishing temporal ordering between the agricultural intervention, depression and psychological stress outcomes. We attempted to minimize this bias by specifying distinct recall periods in the survey, including a six-month period for agricultural intervention exposure, seven-day period for depressive symptoms and one-month period for psychological stress outcomes. However, we were unable to completely rule out reverse causality given the cross-sectional design employed in our study. Future longitudinal studies may be needed to confirm the directionality of the observed association. The generalizability of our study findings is limited to participants enrolled in a group-based microfinance program in Western Kenya. Individual and contextual factors likely influence how such programs are implemented and experienced. Consequently, estimating effects within specific contexts may be more informative than deriving a single estimate that may not apply broadly. Nonetheless, the socioeconomic pathways through which agricultural interventions may influence depression such as improved income stability and food security are likely relevant across similar low-resource contexts, supporting cautious generalizability of our findings to comparable settings.

## Conclusion

The agricultural intervention was associated with modestly lower depression, and psychological stress scores, although overall estimates were imprecise. The intervention appeared most beneficial for individuals with higher levels of depressive symptoms, suggesting agricultural and microfinance support may be particularly beneficial for those experiencing greater psychosocial burden. Integrating agricultural assistance into microfinance programs may help reduce depression, especially among individuals with greater psychological need.

Future longitudinal research with large samples are needed to improve our causal understanding of this relationship, and identify subgroups most likely to benefit.

## Data Availability

All data produced in the present study are available upon reasonable request to the authors

## Acknowledgements

We thank the **B**ridging **I**ncome **G**eneration through grou**P I**ntegrated **C**are (BIGPIC) microfinance members who volunteered to participate in this study.

## Data availability

Data for this research is not publicly available due to participants sensitive information. However, the date may be made available upon reasonable request to the corresponding author.

## Authorship Contribution

Charles Opondo: conceived the research idea, developed the protocol, acquired funding, collected data, performed formal analysis and wrote the original draft; James Kamadi: conceived the intervention idea; design of intervention; project administration, and data collection; James Akiruga Amisi: Writing -review and editing; Sonak Pastakia: conceived the intervention idea; acquired funding; design of intervention; Writing - review and editing; Ricky Camplain: Writing - review and editing, Nana Gletsu-Miller: Writing - review and editing; Christina Ludema: methodology, review, and editing; Molly Rosenberg; supervision, conceptualization, methodology, formal analysis, writing, review, and editing.

---

**Supplementary Table 1:** IPT-weighted linear regression estimates for the association between the agricultural intervention and depression scores, stratified by wealth status.

| | Marginal $\beta$ Coefficient | 95% Confidence Interval | p-value |
| --- | --- | --- | --- |
| **Depression scores** |  |  |  |
| Full Sample N=312 | -1.67 | -3.94, 0.59 | 0.15 |
| Wealth- stratified sample |  |  |  |
| High-wealth<br>N=148 | -0.38 | -4.38, 3.62 | 0.85 |
| Low-wealth<br>N=146 | -5.35 | -10.14, 0.56 | 0.03 |

**Supplementary Figure 1:**
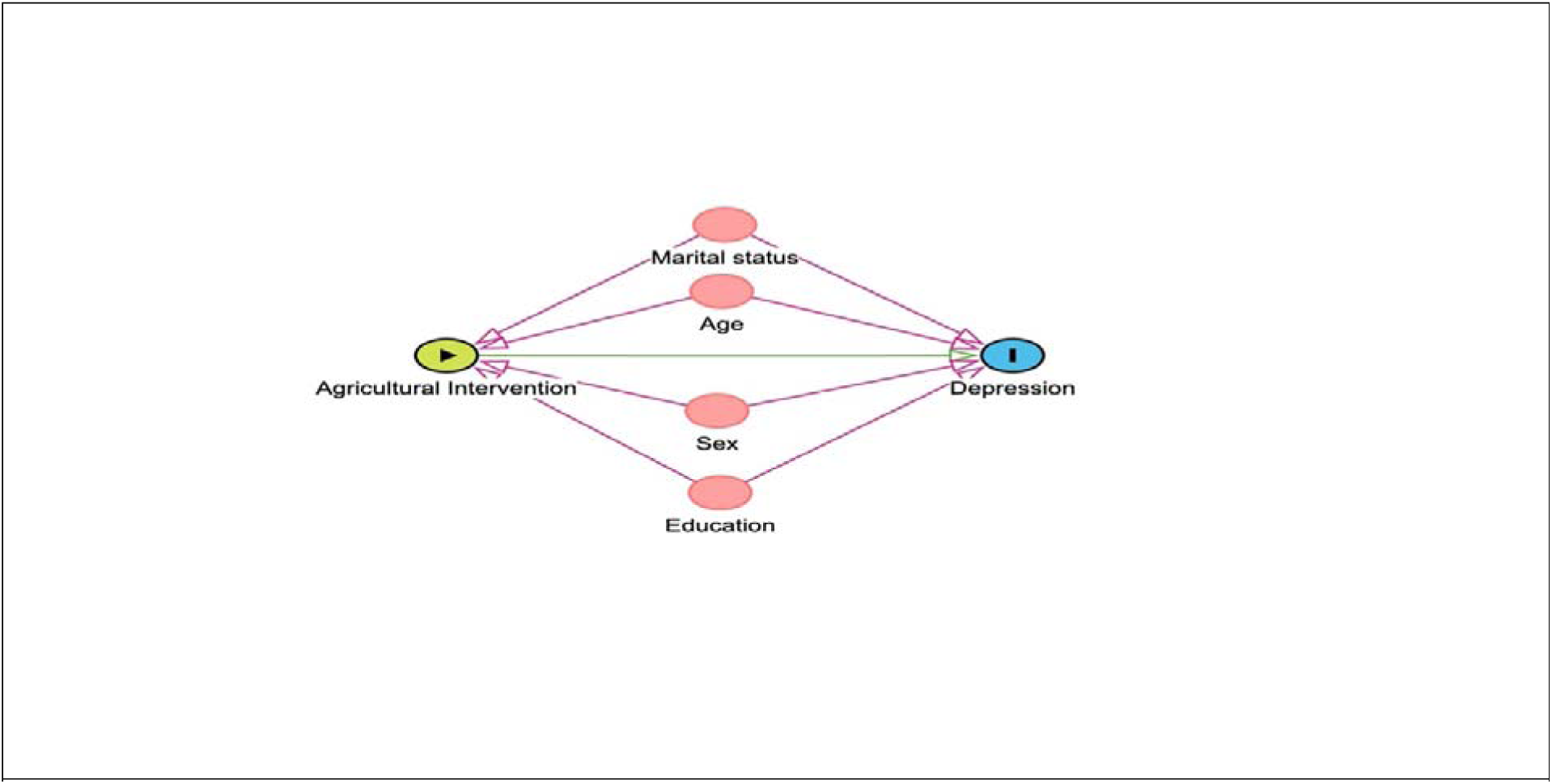
Directed acyclic graph depicting hypothesized relationships between exposure to agricultural intervention and the outcome of depression

